# Chest CT Scan of Hospitalized Patients with COVID-19: A Case-Control Study

**DOI:** 10.1101/2020.04.07.20056762

**Authors:** Lianpin Wu, Qike Jin, Jie Chen, Jiawei He, Jianghu (James) Dong

**Author notes:** Corresponding author: Jianghu (James) Dong, Department of Biostatistics, College of Public Health, University of Nebraska Medical Center, USA.

## Abstract

**Introduction:** This paper sought to investigate the clinical characteristic differences between suspected and confirmed patients with COVID-19 from CT scan to prevent and treat this infectious disease, since the coronavirus outbreak in the world has seriously affected the quality of life.

**Methods:** We proposed to use a retrospective case-control study to give a comparison between suspected patients and confirmed patients in the clinical characteristics.

**Results:** (56%) patients were confirmed for COVID-19 from suspected 167 patients. We find that elder people were more likely to be infected by COVID-19. Among the confirmed 94 patients, 2 (2%) patients were admitted to an intensive care unit, and 0 (0%) patients died during the study period. We find that images of CT scan of patients with a COVID-19 are significantly different from patients without a COVID-19.

**Conclusions:** To our best knowledge, it is the first time to use the case-control design to study the coronavirus disease, since it is particularly appropriate for investigating infectious disease outbreaks. The clinical treatment experience in this study can supply a guideline for treating COVID-19 as the number of the infected patients is increasing in the world. Compared with other studies, we find that the mortality rate and the intensive care unit rate can be reduced if patients can be treated timely in the right identification and detection with nucleic acid testing and chest CT scan. Therefore, we recommend nucleic acid testing and chest CT scan for the clinical treatment practice from this successful clinical treatment study.

## Introduction

In December 2019, many cases of pneumonia have been detected in Wuhan, Hubei Province, China. These patients were with similar clinical manifestations of viral pneumonia, characterized by fever and cough and other nonspecific symptoms, including breathing difficulties, muscle aches, and fatigue. Based on the development characteristics and viral structure of this health emergency, the World Health Organization (WHO) named it as COVID-19 ^1^, which is the seventh member of the human-infected coronavirus family 2, and two other notable family members are severe acute respiratory syndrome (SARS) coronavirus and Middle East Respiratory Syndrome (MERS) coronavirus, which have caused significant public health hazards. The paper ^2^ confirmed that COVID-19 can be transmitted from people to people through droplets or contact, or through the fecal-oral route, with a high incidence and rapid infection, posing a huge threat to global public health. COVID-19 is spreading around the world. The coronavirus outbreak in the world has seriously affected the quality of life.

One main purpose is to show the clinical treatment experience in COVID-19 from CT scan. Patients in this study were different from Wuhan patients in the paper ^3^. All the suspected patients with/without the symptoms were concentrated to the hospital in the fever clinic areas, and then they were carried out by physical examinations including the blood routine test, C-reactive protein test, nucleic acid examination, and chest CT scans. If the abnormal and blood routine did not indicate the obvious signs of bacterial infection, then patients were becoming a suspected case. After that, the new coronavirus nucleic acid examination was carried out to test whether it was positive or negative. Positive patients were further transferred to the special ward for treatment with the chest CT scans, which can show the cloudy lungs on both sides of each infected person. Therefore, this case-control study wants to investigate whether nucleic acid testing and CT imaging examination are the important key to detect the lung infection and identify the suspected COVID-19 infections, since rapid identification of diseases can ensure timely treatment with very limited health resources for a large amount number of sick patients.

To demonstrate the clinical outcomes of the above treatment process, we compared the patient character and clinical treatment outcomes of confirmed patients with those suspected patients. We investigated the progression analysis review of the CT scans. We find that this case-control study has more cases than the initial partial reported study ^4^ with total 62 cases. Therefore, we believed that this retrospective case-control report can provide some detail data for understanding the Coronavirus from the perspective of CT scan results.

## Methods

### Data resource

The 167 consecutive hospitalized patients collected from January 21 to Feb 11, 2020, and the final date of follow-up was February 28, 2020. Patient epidemiological data, clinical symptoms, blood routine test, and chest CT characteristics were collected in the hospital in standard form. The chest CT Scan was performed using GE Light Speed VCT 64-slice and Phillips Brilliance 16-slice CT. The scanning parameters are tube voltage 120 kV, automatic tube current, scanning layer thickness 5.00 mm, reconstruction layer thickness 11.5 mm, pitch 0.99 1.20, image reconstruction using high-resolution algorithm, matrix 512 × 512. Lung window with position 500 HU and window-width 1500 HU and mediastinum window with position 40 HU and window-width 400 HU.

The chest CT scan and image data have covered the comprehensive features for each patient as follows: (1) the location of lesions according to the left, right, or both lungs, (2) the number of lesions, (3) the location of the lesions along the bronchus beam distribution, near pleural distribution, or mixed distribution, (4) the lesion size, (5) the presence of air bronchi sign or not, (6) the presence of grid-like texture, (7) the lesion morphology, (8) lung involvement percentage (9) The presence of atelectasis or not, (10) whether the density of the lesion is ground-glass-like, solid or mixed, (11) the presence of pleural effusion or the presence of lymphadenopathy in extrapulmonary manifestations, and (12) other potential lung diseases features such as pulmonary bullae, pulmonary nodules, calcifications, and fibrous lesions.

### Statistical analysis

It is known that the case-control design is particularly appropriate for investigating infectious disease outbreaks. We divided all suspected patients into the control group and the case group from the nucleic acid testing result. The case group included the suspected patients with a positive nucleic acid testing result, and the control group included patients with a negative nucleic acid testing result. We summarized continuous variables as either means and standard deviations or medians with interquartile ranges for all patients in control and case groups. For categorical variables, we calculated the percentages of patients in each category. Characteristics for case and control patients were compared using analysis of variance or nonparametric statistics (rank sum tests) and Chi-square tests as appropriate. All analyses were done with SAS software, version 9.4.

### Patient and public involvement

No patients were involved in the clinical trial design, setting the research questions, and the outcome measures directly. No patients were asked to advise on interpretation or writing up of results.

## Results

### Patient epidemiological characteristics

The 167 COVID-19 suspected patients were included in the statistical analysis. Their aged 1- 87 and the mean (SD) age of the patients was 44 (19) years. 94 confirmed cases (56%) and 67(45%) control group. 92 (55%) were male and 75 (45%) were females, with men accounting for the largest proportion. In COVID-19 patients, there were 38 cases (23%) in Wuhan. There were 8 cases of hypertension 23 (14%), 13 (8%) cases of diabetes, 13 (8%) cases of Cardiogenic diseases, 3 cases of lung disease (1%), 1 case of kidney disease, and 2 (1%) cases of liver disease. 123 (74%) cases of first symptoms, the most common symptoms are 123 (74%) cases of fever, 105 (63%) cases of cough, 22 (13%) cases of runny nose, 22 (13%) cases of gastrointestinal symptoms, 42 (25%) cases of sore throat, 32 (19%) cases of fatigue, 31 (19%) cases of muscle pain. 47% patients have been treated by Chinese medicine, and it may help fight the COVID-19 disease. The summary of basic characteristics in all 167 patients with COVID-19 is shown in Table 1.

**Table 1:** Patient epidemiological characteristics.

|  | Total<br>(N=167) | Positive<br>(N=94) | Negative<br>(N=73) | P- value |
| --- | --- | --- | --- | --- |
| Mean (SD) of Age | 44(19) | 47(14) | 38(23) | <0.001 |
| Categories (%) |  |  |  |  |
| <18 | 11 | 1 | 23 |  |
| 18-39 | 27 | 24 | 30 |  |
| 40-59 | 45 | 57 | 29 |  |
| >=60 | 17 | 18 | 18 |  |
| Male (%) | 55 | 60 | 50 | 0.215 |
| Contact history of Wuhan (%) | 23 | 28 | 17 | 0.041 |
| Comorbidities (%) |  |  |  |  |
| High blood pressure | 14 | 16 | 13 | 0.785 |
| Diabetes. | 8 | 12 | 6 | 0.262 |
| Cardiogenic diseases | 8 | 6 | 8 | 0.881 |
| Lung disease. | 1 | 1 | 0 | NA |
| Anemic. | 0 | 0 | 0 | NA |
| Stroke. | 1 | 1 | 0 | NA |
| Kidney disease | 0 | 0 | 0 | NA |
| Surgery | 11 | 13 | 7 | 0.940 |
| Liver disease. | 0 | 0 | 0 | NA |
| First symptoms including wheezing and coughing (%) |  |  |  |  |
| Fever | 74 | 74 | 75 | 0.647 |
| Cough | 63 | 73 | 55 | 0.606 |
| Runny nose | 13 | 9 | 20 | 0.234 |
| Gastrointestinal symptoms | 13 | 14 | 32 | 0.998 |
| Sore throat | 25 | 19 | 25 | 0.475 |
| Fatigue | 19 | 18 | 18 | 0.998 |
| Muscle pain | 19 | 26 | 14 | 0.078 |
| Body temperature | 37.18(0.79) | 37.03(0.82) | 37.27(0.78) | 0.072 |
| Pulse | 90(19) | 84(14) | 92(20) | 0.003 |
| Respiratory rate | 19.76 | 19.1(1.77) | 20.2(3.27) | 0.005 |
| Blood pressure | 42(2.13) | 47(14) | 38(23) | <0.001 |
| Medicine (%) |  |  |  |  |
| Antivirus | 73 | 75 | 71 | 0.640 |
| Antibiotics | 55 | 45 | 68 | 0.002 |
| Hormone | 13 | 22 | 0 | <0.001 |
| Chinese Medicine | 26 | 47 | 0 | <0.001 |

### Biochemical testing

The results of blood routine tests of 167 patients were summarized in Table 2, where the confirmed patients were compared with patients with the negative result from nucleic acid testing.

**Table 2:** The results of blood routine tests from clinical blood laboratory.

|  | Total<br>(N=167) | Positive<br>(N=94) | Negative<br>(N=73) | P- value |
| --- | --- | --- | --- | --- |
| Median (IQR) of C-reactive protein | 7.10<br>(1.30,17.50) | 8.25<br>(1.57,15.40) | 6.75<br>(1.07, 30.19) | <0.001 |
| Median (IQR) of White blood cell count | 5.38<br>(4.45,7.67) | 4.89<br>(4.05,5.68) | 7.99<br>(5.08,10.03) | <0.001 |
| Median (IQR) of Red blood cell count | 4.65<br>(4.09,5.00) | 4.49<br>(4.03,4.90) | 4.78<br>(4.40,5.08) | 0.002 |
| Median (IQR) of Lymphocyte count | 1.16<br>(0.93,1.56) | 1.30<br>(0.96,1.51) | 1.11<br>(0.77,1.66) | <0.001 |
| Median (IQR) of Hemoglobin. | 137.24<br>(109.00,149.00) | 134.20<br>(108.11,149.23) | 139.00<br>(114.00,150.00) | 0.864 |
| Median (IQR) of Platelet count. | 142.00<br>(59.00,185.00) | 145.1<br>(65.21,180.23) | 134.00<br>(46.00,198.00) | 0.166 |
| Median (IQR) of Plasma prothrombin time determination | 13.65<br>(13.00,13.90) | 13.50<br>(13.10,13.81) | 13.80<br>(10.80,15.15) | <0.001 |
| Median (IQR) of Thrombin time | 15.21<br>(14.45,15.76) | 15.10<br>(14.43,15.70) | 15.50<br>(14.50,16.10) | <0.001 |
| Median (IQR) of Activated partial thromboplastin. | 41.70<br>(40.00,45.80) | 43.46<br>(40.10,45.60) | 41.30<br>(38.00,46.80) | 0.003 |
| Median (IQR) of D-D dimer mg/liter | 0.45<br>(0.30,0.70) | 0.40<br>(0.08, 0.60) | 0.70<br>(0.44,1.03) | 0.416 |
| Median (IQR) of Alanine aminotransferase | 33.00<br>(8.00,45.00) | 33.00<br>(7.00, 40.00) | 25.50<br>(8.00, 53.00) | 0.021 |
| Median (IQR) of Urea nitrogen | 3.97<br>(3.39,4.90) | 4.03<br>(3.43, 4.66) | 3.76<br>(3.15, 6.19) | 0.001 |
| Median (IQR) of Fibrinogen. | 4.90<br>(4.06,5.67) | 4.88<br>(4.07, 5.67) | 5.03<br>(3.31, 5.90) | 0.002 |
| Median (IQR) of Eosinophil count | 0.03<br>(0.01,0.09) | 0.02<br>(0.01, 0.07) | 0.04<br>(0.01, 0.11) | <0.001 |
| Median (IQR) of Hematocrit | 0.40<br>(0.37,0.44) | 0.39<br>(0.36, 0.42) | 0.41<br>(0.38, 0.45) | 0.005 |

### Chest CT scan and image

In the first chest CT scan of 167 COVID-19 patients, 85% confirmed patients have lesions in both of left and right lung compared 30% in suspected patients. 78% confirmed patients had more than 3 lesions. 95% confirmed patients were with patch lesion morphology. The detail comparative results are summarized in Table 3.

**Table 3:**
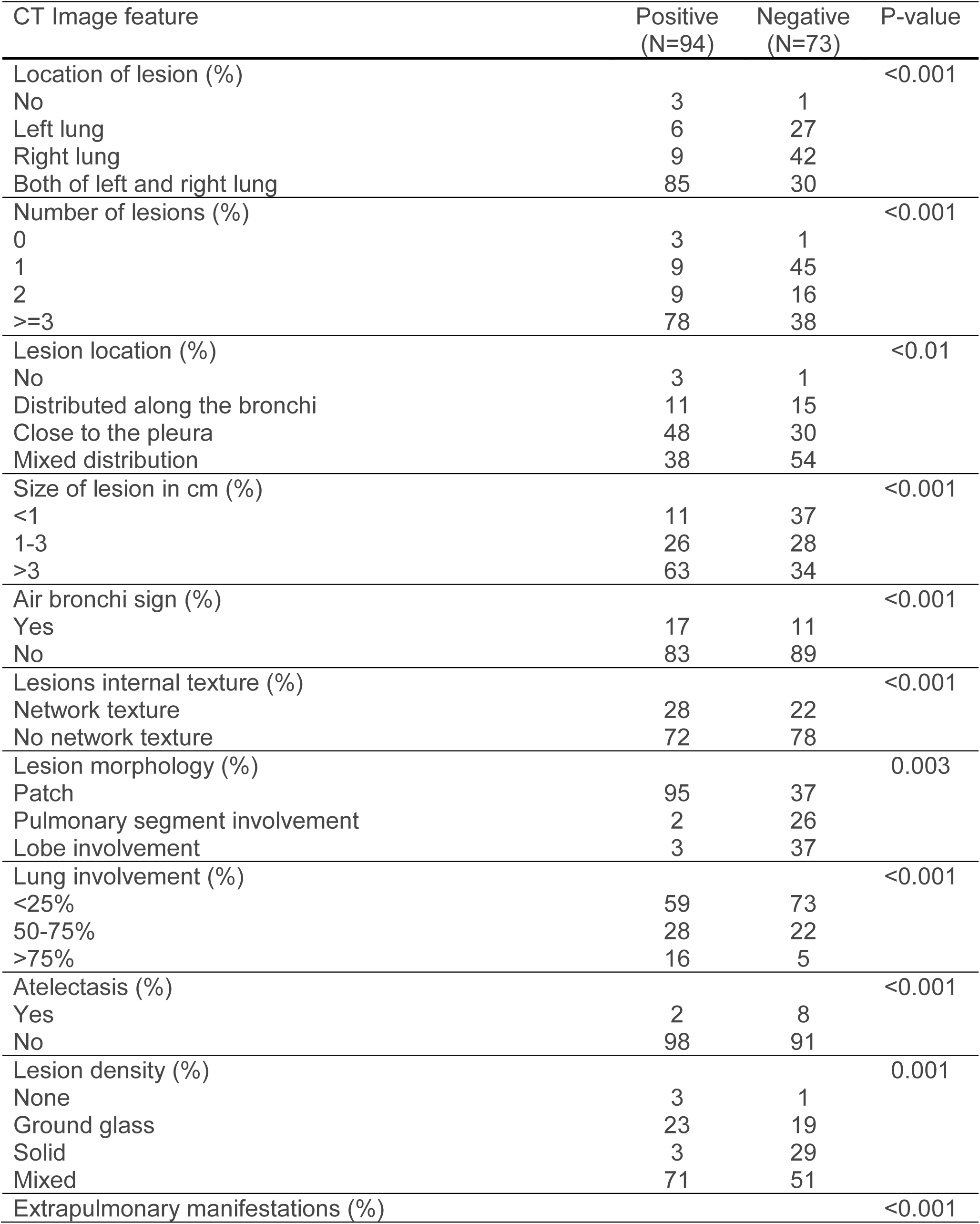
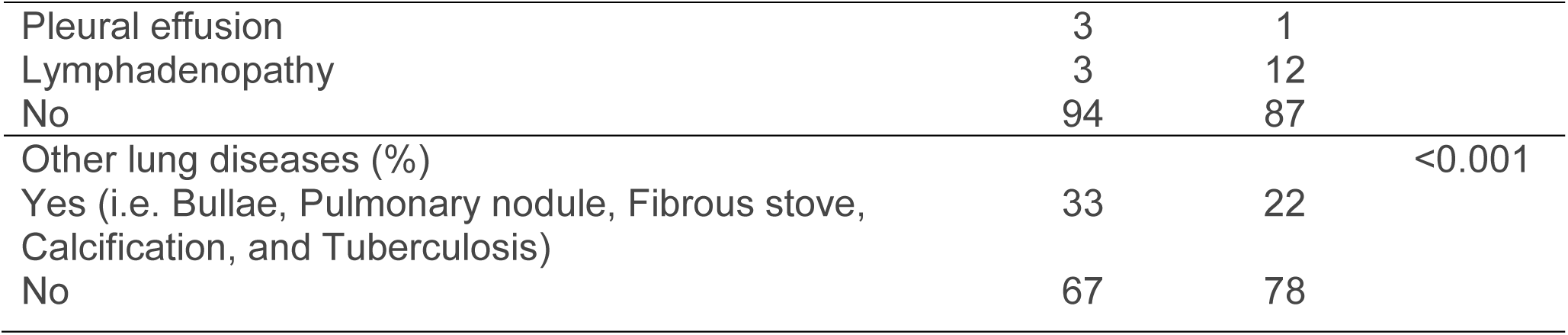
Chest CT results with lesions and imaging manifestations.

CT scans of eight random selected patients were shown in Figure 1. We find that CT scans of patients with a coronavirus are significantly different from patients without a coronavirus infection. For example, the lesions are multiple patchy, and the grinding glass shadows are distributed in the peripheral pulmonary field in the infected patients. The lesions are large, strip-like, uneven density, and to the lung leaf or lung section distribution, the neighbor’s pleural reactive slight thickening in patients without a virus infection.

**Figure 1:**
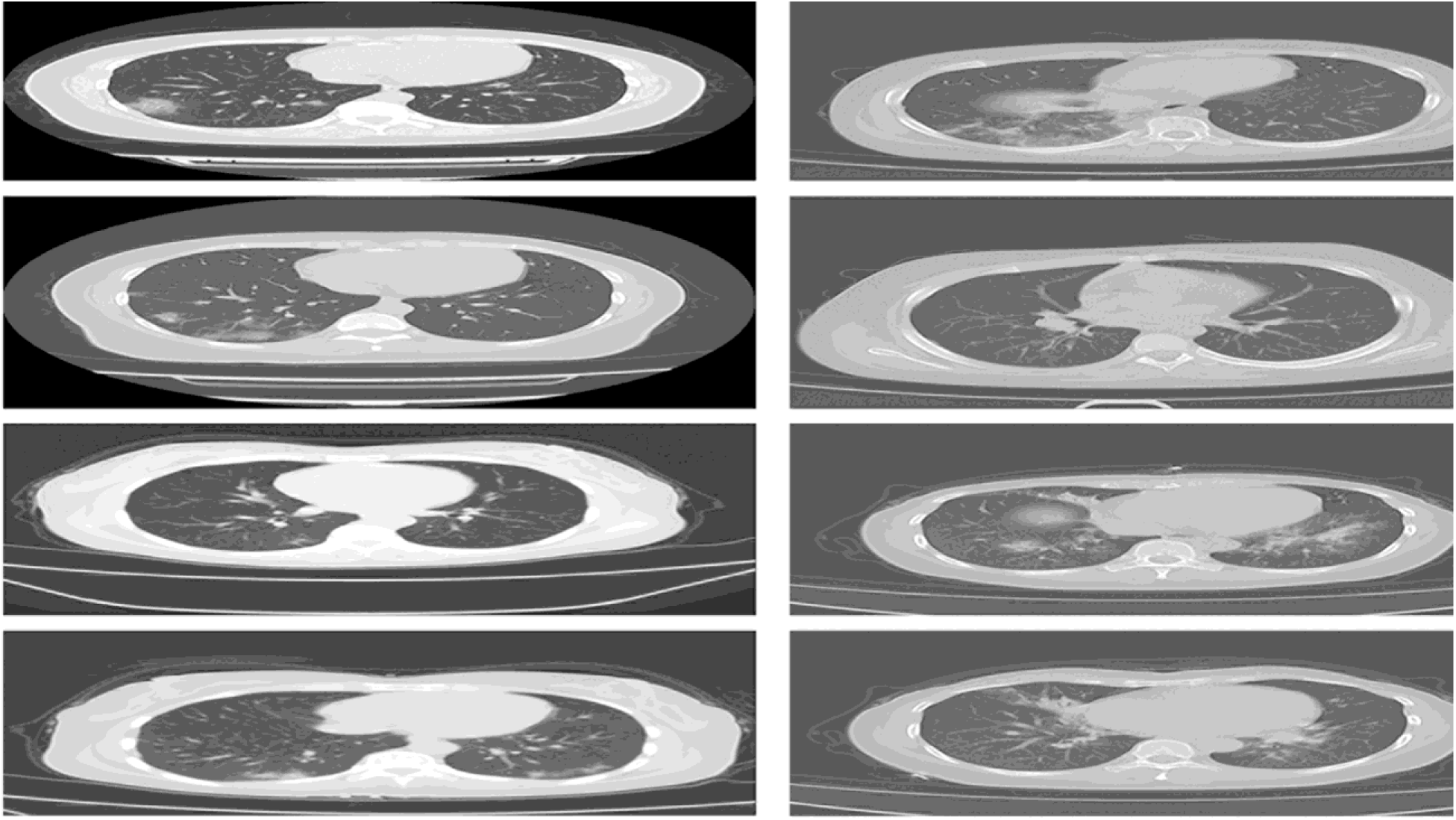
Left panel are CT scans of four random selected patients with a positive nucleic acid testing. Their lesions are multiple patchy, and the grinding glass shadows are distributed in the peripheral pulmonary field. Right panel are CT scans of four random selected patients with a negative nucleic acid testing. Their lesions are large, strip-like, uneven density, and to the lung leaf or lung section distribution, the neighbor’s pleural reactive slight thickening.

### Intensive Care Unit

Two (2%) patients in the case group were admitted to an intensive care unit (ICU) in this study. It is different from the recent report ^5^ with 26% patients in ICU. Both of ICU patients were elder patients with hypertension or diabetes. 0 (0%) patients died during the followed-up period.

## Conclusions

This paper gave the clinical characteristic differences between suspected and confirmed patients with COVID-19. 94 (56%) COVID-19 patients were identified from the suspected 167 patients in this case-control study. We find that fever symptoms account for the first clinical characteristics since 74% of patients had a fever/cough symptom, and then followed other gastrointestinal reactions, sore throat, fatigue, and muscle pain. Laboratory examination showed normal or decreased peripheral white blood cells, decreased lymphocyte count, and increased C-reactive protein (CRP) level. Their clinical characteristics did not fully cover all patients such as first symptoms. We also find that some suspected patients need to be combined with epidemiological history since the first nucleic acid testing were negative, but they were confirmed positive later. Elder people were more likely to be infected by COVID-19 from Table 1. Among the confirmed 94 patients, 2 (2%) patients were admitted to an intensive care unit, and 0 (0%) patients died during the study period. Compared with the clinical result in the paper ^6,7^, our clinical treatments were more successful from the clinical treatment process.

The strengths of this case-control design can clearly show the difference between the confirmed and suspected patients. The comparative results can provide some valuable information in the understanding of the clinical features of COVID-19 and identifying the potential patients from suspected patients, especially in the current period when the number of people with the suspected disease is increasing rapidly in the world. We recommend that the nucleic acid examination and chest CT scan are important tools to detect COVID-19 and stop the deterioration process of the lung function to Intensive Care Unit from our clinical treatment experience.

## Data Availability

Homepage: www.unmc.edu/publichealth/departments/biostatistics/jianghu-dong.html

## ICMJE statement

All authors meet the ICMJE authorship criteria

## Conflicts of interest

None

## Ethics statement

Ethics approval for this project was obtained from the Institutional Review Board of the University of Nebraska Medical Center (1982933).

## Notes

### Competing Interest Statement

The authors have declared no competing interest.

